# Foetal Allogeneic Intracerebroventricular Neural Stem Cell Transplantation in People with Secondary Progressive Multiple Sclerosis: A phase I dose-escalation clinical trial

**DOI:** 10.1101/2022.11.14.22282124

**Authors:** MA Leone, M Gelati, DC Profico, C Conti, C Spera, G Muzi, V Grespi, I Bicchi, C Ricciolini, D Ferrari, M Zarrelli, L Amoruso, G Placentino, P Crociani, F Apollo, P Di Viesti, D Fogli, T Popolizio, C Colosimo, D Frondizi, G Stipa, E Tinella, A Ciampini, S Sabatini, F Paci, G Silveri, C Gobbi, E Pravatà, E Zecca, RF Balzano, J Kuhle, M Copetti, A Fontana, M Carella, G D’Aloisio, L Abate, Y Ventura Carmenate, S Pluchino, L Peruzzotti-Jametti, AL Vescovi

## Abstract

**Background:** Advanced cell therapeutics are emerging as potentially effective treatments for chronic neurological diseases, including secondary progressive multiple sclerosis (SPMS). Here we report the results of a phase I trial in which good manufacturing practice-grade foetal allogeneic human neural stem cells (hNSCs) were implanted via intracerebroventricular (ICV) injection in 15 individuals with active and non-active SPMS.

**Methods:** This is a phase I, open-label, multicentre, dose-escalation, international study. The primary objective was to assess the feasibility, safety, and tolerability of ICV injections of allogeneic hNSCs in patients affected by SPMS over a study follow up of 12 months. We also evaluated the number and type of adverse events (AEs) leading to a maximum tolerated dose, the general health status, and mortality. The secondary objectives were the therapeutic benefit of allogeneic hNSCs using assessment scales, magnetic resonance imaging (MRI), and laboratory and neurophysiologic parameters.

**Findings:** Fifteen unrelated SPMS patients were enrolled and treated between 2018 and 2020. The participants had a median age of 49.8 years. Their mean extended disability status scale (EDSS) at enrolment was 7.6, the mean disease duration was 22 years, and mean time from diagnosis to progression was 10.1 years. Neither treatment-related deaths nor serious AEs were reported during the study (1 year follow up after treatment). All the other AEs were classified as non-serious and were associated to non-study concomitant therapy or other medical conditions not connected to the experimental treatment. During the study, none of the participants worsened in the progression of their SPMS as shown by the evaluation scales implemented to assess their progress. Laboratory and neurophysiologic parameters showed no clinically significant variations. MRI follow-up showed non-clinically significant type 1, 2, and 3 changes.

**Interpretation:** The intracerebroventricular injection of foetal allogeneic hNSCs in people with SPMS is feasible, tolerated and safe. Study participants displayed a substantial clinical stability during the 12-month follow-up. The absence of relevant adverse reactions (Ars) arising from the transplantation of hNSCs indicates a short-term neutral balance between benefits and risks and suggests a concrete, though perspective therapeutic possibility for SPMS patients. Further studies are needed to confirm and extend the findings herein and evaluate the actual therapeutic potential of advanced cell therapeutics for a condition where the lack of effective disease modifying therapies is a major unmet clinical need.

## INTRODUCTION

Multiple sclerosis (MS) is the most common cause of non-traumatic disability in young adults, with over 2·5 million sufferers worldwide.^1^ The last decade has seen a fast development of effective disease modifying treatments (DMTs) for patients with a relapsing remitting form of MS (RRMS). However, as the disease evolves into secondary progressive MS (SPMS), DMTs have limited efficacy. Thus, SPMS patients currently have a major unmet need.^2,3^ A solution to the current lack of effective treatments for SPMS may come from emerging cell therapy approaches, which have shown promising initial results in other central nervous system (CNS) diseases, such as amyotrophic lateral sclerosis (ALS) and Parkinson’s disease.^4^

*In vivo* studies in rodent and non-human primate models of MS have shown that human neural stem cells (hNSCs)^5-8^ are an effective and safe tool to induce CNS functional recovery due to their tissue specificity and multiple mechanisms of action.^9,10^ hNSCs not only repair the damaged CNS by replacing cells lost to injury, but also exert immunomodulatory actions on both innate and adaptive immune responses via secretion of trophic factors, cross-correction of missing enzymatic activities, and metabolic reprogramming.^11-14^ Additional data suggest that delivering these cells directly to the CNS via a single intracerebroventricular (ICV) injection allows maximising the number of cells that reach the CNS^15-17^ and may be key to target the intense compartmentalised inflammation that drives progression in SPMS.^18^ Indeed, following ICV injection, transplanted hNSCs spread throughout the ventricular and subarachnoid space,^16,19^ enabling their inflammation-guided migration into the CNS, where they may reach axons and myelinating cells directly without crossing the blood brain barrier.^20^ The ICV cell injection is based on a widely used, technically simple, rapid, and standardised neurosurgical procedure with a minimal rate of complications (6·8%), mainly due to catheter malposition, haemorrhage, and infection (Morgenstern et al., 2016). ICV is also a relatively standardised experimental procedure, since it has already been used for the injection of growth factors in ALS patients^21^ and for chemotherapeutic agents in anti-tumour therapy. ^22^

Here, we report the results of the first phase I, open-label, multicentre, dose-escalation study in which good manufacturing practice (GMP)-grade foetal allogeneic human neural stem cells (hNSCs) were implanted via intracerebroventricular (ICV) injection in 15 individuals with SPMS. To the best of our knowledge, this is also the largest phase I trial with hNSCs in people with SPMS conducted to date.

## METHODS

### Study design

In this phase I, open-label, multicentre, international, dose-escalation study, the participating patients were affected by SPMS with progressive accumulation of disability after an initial relapsing course, with or without disease activity. Fifteen patients, between 18 and 60 years of age were enrolled according to a “standard” dose-escalation phase I design following a modified Fibonacci sequence (100%, 60% and 50% dose increments). After the initial screening, all screened patients entered a 3-month run-in phase. After that, the patients were prospectively enrolled into four cohorts receiving four different doses of allogenic hNSCs (5, 10, 16 and 24 million cells).

This study was performed in three participating centres located in Italy and Switzerland. The Italian centres, the “IRCCS Casa Sollievo della Sofferenza” Research Hospital (Site 1) and the “Santa Maria di Terni” Hospital (Site 2), recruited the patients. The MSC of the Neurocentre of Southern Switzerland performed the magnetic resonance imaging (MRI) analysis, while ICV treatment was performed at Site 2.

The study was approved by the Ethical Committee of the Istituto Tumori “Giovanni Paolo II” (Bari) from the “Fondazione IRCCS Casa Sollievo della Sofferenza” Research Hospital (01PU/2016–21-01-2016); the Ethical Committee of the “Aziende Sanitarie dell’Umbria” (2404/17); the Agenzia Italiana del Farmaco (AIFA); the Istituto Superiore di Sanità (3090(16)-PRE21-1408–06-04-2016). The trial was subsequently registered in the European Clinical Trials Database (EudraCT, 2015-004855-37), and in ClinicalTrials.gov (NCT03282760).

### Participants

Eligible patients were adults of either sex with a diagnosis of SPMS, with or without disease activity (Lublin 2014) with an Expanded Disability Status Score (EDSS)^23^ ≥ 6.5 and ≤ 8, showing a progressive accumulation of disability after initial relapsing course over the 2 years before recruitment (≥ 1.0 point for patients with EDSS =6.5 at the time of inclusion, and ≥ 0.5 points for patients with EDSS > 6.5 at the time of inclusion), and ineligibility to other therapeutic alternatives (as assessed by the treating neurologist). All patients signed a written informed consent to be enrolled in the study. Exclusion criteria included: other neurologic conditions; psychiatric/personality disorders or severe cognitive decline; history of significant systemic, infectious, oncologic, or metabolic disorders; other autoimmune diseases; chronic infections (HBV, HCV, HIV, tuberculosis); inability to undergo MRI scans; inability to provide informed consent, received immunomodulant/immunosuppressive treatments <6 months before inclusion; participated in other research; any contra-indication to lumbar puncture; and were pregnant or breast feeding.

At the end of the run-in period, and if no serious co-morbidity nor health status changes occurred, patients were deemed eligible for the intervention. After eligibility assessment and provision of informed consent, each eligible patient was registered in the “Database for Clinical Studies with Gene and Somatic Therapy” of the “Istituto Superiore di Sanità” (ISS). Before registration, the ISS verified patient eligibility to ensure criteria were respected. Upon registration, the database assigned a unique identification number to each patient which was used for anonymisation purposes.

To establish baseline clinical features, a 3-month run-in period was started after the screening examination. All patients were evaluated at the onset and the end of the run-in period by investigating and performing physical and neurological examination, vital signs, pregnancy test in fertile women, haematological and urine tests, lumbar puncture for standard CSF examination and JC Virus test, serum and CSF collection (stored at minus 80 degrees), motor, sensory and visual evoked potentials, optical coherence tomography (OCT), EDSS, MFSC, RAO brief repeatable battery, MS-QOL54 for the evaluation of quality of life (Solari et al. 1999), brain and spinal MRI. At the end of the run-in period, and if no serious co-morbidity nor health status changes occurred, patients were deemed eligible for the intervention. Around 50% of patients had at least one new or one enlarging T2-visibile lesion, while 43% of patients had at least one lesion with contrast enhancement during the run-in period.

### Study objectives and outcomes

The primary objective of the study was to assess the feasibility, safety, and tolerability of foetal allogeneic hNSCs delivered via ICV injection in people with active and non-active SPMS patients by evaluating the following outcomes: mortality, the number and type of adverse reactions (ARs) or events (AEs) leading to a maximum tolerated dose.

The secondary objectives were to evaluate the functional effects of hNSC therapetics by monitoring the following outcomes of disease progression: functional disability via EDSS and Multiple Sclerosis Functional Composite (MSFC)^24^ ; annualised relapse rate and time to confirmed relapse; cognitive function by Rao’s brief repeatable battery [BRB] of neuropsychological tests; visual, sensory and motor functions via combination of electrophysiological measurements (visual, somato-sensory, motor evoked potentials [EP]); and optical coherence tomography (OCT). Brain MRI evaluations were performed to monitor structural changes related to both the intervention and disease activity. MRIs were acquired at the two recruiting centres, using a Philips Ingenia scanner (Philips Medical Systems, Best, The Netherlands) and a Siemens Verio scanner (Siemens, Erlangen, Germany). Sequences type and parameters were harmonised between the two vendors to improve reproducibility. The MRIs were performed at run-in onset (month -3), run-in end (month 0), and at months +1, +2, +3, +4, +5, +6, +9 and +12.

Potential effects of the treatment on biomarkers of neuronal loss and inflammation was also evaluated. Cerebrospinal fluid (CSF) and serum neurofilament (NfL) levels were used as markers of neurodegeneration/neuronal damage, while CHI3L1 (also known as YKL-40) as indicator of reactive astrocytes.^25^ We also investigated serum and CSF levels of IL-17a, IL-2, IL-8, TNF-α, CCL2, CCL3, CX3CL1, VEGF-a, OPN and GFAP as additional exploratory objectives. All the biomarkers were evaluated in both CSF and serum collected before and after treatment (see table 1 for time points) Clinical evaluation

**Table 1:** Clinical trial study plan

|  | Screening | Run in<br>START | Run in<br>END | Surgery | M1 | M2 | M3 | M4 | M5 | M6 | M7 | M8 | M9 | M10 | M11 | M12 |
| --- | --- | --- | --- | --- | --- | --- | --- | --- | --- | --- | --- | --- | --- | --- | --- | --- |
| Informed consent signature | x | x |  |  |  |  |  |  |  |  |  |  |  |  |  |  |
| Medical history | x |  |  |  |  |  |  |  |  |  |  |  |  |  |  |  |
| Neurological history | x |  |  |  |  |  |  |  |  |  |  |  |  |  |  |  |
| Physical examination | x | x | x |  |  |  |  |  |  |  |  |  |  |  |  |  |
| Infectious screening | x |  |  |  |  |  |  |  |  |  |  |  |  |  |  |  |
| Vital Signs | x | x | x |  | x |  | x |  |  | x |  |  | x |  |  | x |
| Pregnancy test for fertile women | x | x | x |  |  |  |  |  |  |  |  |  |  |  |  |  |
| Hematological tests | x | x | x |  | x | x | x | x | x | x | x | x | x | x | x | x |
| Electrocardiogram | x |  |  |  |  |  |  |  |  |  |  |  |  |  |  |  |
| Chest X-ray | x |  |  |  |  |  |  |  |  |  |  |  |  |  |  |  |
| standard urine test | x | x | x |  | x |  | x |  |  | x |  |  | x |  |  | x |
| Neurological examination | x | x | x |  | x | x | x | x | x | x | x | x | x | x | x | x |
| VAS |  | x | x |  | x | x | x | x | x | x | x | x | x | x | x | x |
| EDSS |  | x | x |  | x |  | x |  |  | x |  |  | x |  |  | x |
| MSFC |  | x | x |  | x |  | x |  |  | x |  |  | x |  |  | x |
| Evoked potential test |  | x | x |  | x |  | x |  |  | x |  |  | x |  |  | x |
| OCT |  | x | x |  | x |  | x |  |  | x |  |  | x |  |  | x |
| Brain and spinal MRI |  | x | x | x | x | x | x | x | x | x |  |  | x |  |  | x |
| RAO battery |  | x | x |  | x |  | x |  |  | x |  |  | x |  |  | x |
| MS-QOL54 |  | x | x |  | x |  | x |  |  | x |  |  | x |  |  | x |
| Conselling with psychologist | x |  |  |  | x |  |  |  |  |  |  |  |  |  |  |  |
| Serum collection for biomarkers |  | x | x |  | x |  | x |  |  | x |  |  | x |  |  | x |
| CSF collection for biomarkers |  | x | x |  | x |  |  |  |  | x |  |  |  |  |  | x |
| JC virus examination |  | x | x |  | x |  |  |  |  | x |  |  |  |  |  | x |
| AE record |  |  |  | x |  | x | x | x | x | x | x | x | x | x | x | x |
| Thin slice cranial CT or MRI |  |  |  | x | x |  |  |  |  |  |  |  |  |  |  |  |

After ICV injection, the participants were followed up for 12 months post-treatment (**Table 1**). General health status and mortality evaluations were based on the occurrence of any serious co-morbidities or changes in the general health of the participants post-intervention and the number of deaths due to the treatment, or the procedure itself.

All AEs were recorded and evaluated for their relationship with the hNSCs or injection procedure. Each AE was classified and categorised according to the International Conference on Harmonisation guidance for Clinical Safety Data Management’s Definitions and Standards for Expedited Reporting E2A and the Common Terminology Criteria for Adverse Events (CTCAE). Clinical relapses were defined as the appearance of a new neurologic deficit, or worsening of previously stable or improving pre-existing neurologic deficit, separated by at least 30 days from the onset of a preceding clinical demyelinating event.^26^ The neurologic deficit must have been present for at least 24 hours and occurred in the absence of fever (<37 ·5°C) or known infection.

### hNSCs dosage and production

hNSCs were produced following a previously described method^27^ and in full compliance with the conditions and practices required by GMP regulations. The hNSCs consisted of a highly enriched population of cells extracted from a single female foetal human donor (spontaneous miscarriage 16weeks after conception) under Ethics Committee approval, with the donors’ parents’ informed consent, and according to the Helsinki declaration. hNSCs were cultured for 10-17 passages *in vitro*.^27^, the last passage was performed 24-96 hours before formulation of the final drug product.

On the day of treatment, hNSCs were collected from culture flasks, centrifuged counted and suspended in HBSS at a concentration of 50,000 cells/µl. After batch release, the cells were maintained at 4 +/-2 °C for up to 1·5 hours prior to implantation. See Profico et al 2022 for complete quality control strategy for batch release.

### Intervention

Trial participants were admitted to the Neurosurgery Unit of the “Azienda Ospedaliera Santa Maria” (Terni, Italy) for the ICV injection of hNSCs. Before the intervention, they underwent thin-slice cranial CT or MRI scans to evaluate the ventricular system and plan the procedure. Image guidance was provided by the frameless stereotactic AxiEM system (Stealth station AxiEM electromagnetic tracking system, Medtronic navigation, Louisville, CO, USA) to perform the ventricular cannulation. The correct placement of the catheter was verified based on the egress of CSF. Then, a Rickham reservoir was connected to the ventricular catheter. The participants were then prospectively assigned to receive 5 (n=3), 10 (n=3), 16 (n=3), and 24 (n=6) × 10^6^ cells ICV. All the participants underwent a post-operative CT scan within 24 hours to evaluate any complications.

All the participants received methylprednisolone orally 125 mg 2 hours pre-intervention, and cefazolin 1g IV immediately before and after the hNSCs injection. The immunosuppressive treatment also included oral prednisone with a 28-day taper (consisting of a dose change per week, from 60, 40, 20, to 10 mg q.d). The participants also received Tacrolimus (0·05 mg/kg, oral, b.i.d.), 12 hours after the intervention and then every 12 hours, for 6 months. This drug was titrated to maintain blood levels ranging 5–10 ng/ml.

### Data management and study monitoring

All trial demographic and clinical data was collected by designated investigators at the screening, run-in, and follow-up visits using ad-hoc case report forms (CRFs). A clinical trial monitor periodically reviewed the CRFs with source data verification and requested corrections as needed. The data were then entered into a eCRF and underwent systematic quality control of the study manager prior to database locking. The database was periodically reviewed by an independent Data and Safety Monitoring Board (DSMB) to ensure protocol adherence and to monitor possible AEs. The DSMB made recommendations concerning the continuation, modification, or termination of the trial as needed.

### Statistical Methods

Demographic, clinical, and laboratory patients’ characteristics were reported as median and interquartile ranges (IQR) or as mean and standard deviation (SD), as appropriate, for continuous variables and as frequency and percentage for categorical variables. Normal distribution was checked using the Shapiro-Wilk test. Safety analyses were performed in all subjects receiving at least one injection of hNSCs. All AEs and severe AEs (SAEs) were recorded at follow-up visits and at the end of the study. Exploratory efficacy analyses were conducted fashion in all subjects receiving at least one injection of hNSCs (here, FAS and ITT populations matched). Because of small sample size and of almost all variables having non-normal distributions, pre-post differences (12-month visit vs run-in end visit) were assessed via linear models using ranks. New or enlarging T2-visible lesions and lesions with contrast enhancement were analysed using negative binomial models with follow-up time as the offset and results reported as annualised rates.

A p-value <0.05 was considered statistically significant. All analyses have been performed using SAS Release 9.4, SAS Institute, Cary, NC, USA.

## RESULTS

### Clinical Results

A total of 220 candidates applied to participate in this study between September 26, 2017, and January 13, 2020. After undergoing the screening visit and completing the run-in period, fifteen SPMS patients were enrolled in the study. The participants had a median age of 49.8 years (range: 37.8–56.6), eight of them were females and seven were males (1.14 : 1 ratio), and an almost equal proportion was recruited at the two study sites. The mean EDSS was 7.6 (range 7–8), mean disease duration was 22 years (range 16–29), and mean time from diagnosis to progression was 10.1 years (range: 1–20). All the patients’ detailed demographic and clinical characteristics are reported in **Table 2**.

**Table 2:** Demographic data and group assignment.

| DOSE | Gender | EDSS | AGE AT<br>TREATMENT<br>(range) |
| --- | --- | --- | --- |
| 5*10<sup>6</sup> | M | 7,5 | 55-59 |
|  | M | 7 | 50-54 |
|  | M | 8 | 50-54 |
| 10*10<sup>6</sup> | M | 7,5 | 36-39 |
|  | F | 7 | 50-54 |
|  | F | 8 | 50-54 |
| 16*10<sup>6</sup> | F | 7 | 50-54 |
|  | F | 8 | 50-54 |
|  | F | 8 | 45-49 |
| 24*10<sup>6</sup> | F | 7,5 | 50-54 |
|  | F | 8 | 45-49 |
|  | M | 7 | 40-45 |
|  | F | 7 | 55-59 |
|  | M | 8 | 36-39 |
|  | F | 8 | 50-54 |

No deaths nor serious ARs were reported during the study period. The AEs that occurred, including their severity, relationship to study and non-study therapies, actions taken, and outcomes are reported in **Table 3**.

**Table 3:** Adverse events

| Patient | Description of event | expected? | Severity | Relationship to study therapy | Action taken | If other, please specify | New treatment /therapy given/taken? | Outcome (at the end of the study) |
| --- | --- | --- | --- | --- | --- | --- | --- | --- |
| #1 | Tremor | Yes | Mild | Unrelated | Tacrolimus dose changed |  | No | Recovered |
|  | Flu like syndrome | No | Mild | Unrelated | None |  | Yes | Recovered |
|  | Respiratory failure | Yes | Mild | Unrelated | Concomitant therapy changed |  | Yes | Recovered |
|  | Cataract (left eye; surgical procedure) | No | Mild | Unrelated | None |  | No | Recovered |
|  | Upper respiratory infection | No | Mild | Unrelated | Concomitant therapy changed |  | Yes | Recovered |
| #2 | Hyperglycemia | Yes | Moderate | Unrelated | Concomitant therapy changed |  | No | Recovered |
|  | Urinary retention | Yes | Moderate | Unrelated | Other, specify | Catheterization | Yes | Recovering (improving) |
|  | Back pain | No | Mild | Unrelated | Other, specify | New therapy given | Yes | Recovered |
| #4 | Seizure | Yes | Mild | Possible | Other, specify | Diazepam, levetiracetam | Yes | Recovered |
| #7 | Psychosis | No | Moderate | Unrelated | Concomitant therapy changed |  | Yes | Recovered |
| #8 | Fracture (right femur) | No | Moderate | Unrelated | Other, specify | Follow-up 10 omitted | No | Recovered with sequelae |
| #9 | Leukoencephalopathy | No | Mild | Unrelated | Other, specify | MRI evaluations | No | Recovering (improving) |
|  | Urinary tract infection | No | Mild | Unrelated | Other, specify | Drugs administration | Yes | Recovered |
| #12 | Depression | No | Moderate | Unrelated | None |  | No | Unknown |
| #14 | Urinary tract infection | No | Mild | Unrelated | Other, specify | Antibiotics administration | Yes | Not recovered |

Among these, SAEs (defined as AEs requiring hospitalisation) occurred in 2 patients (2/15), but none of them were related to the hNSC injection: of these two, one patient developed a steroid-induced acute psychiatric disorder 1 month post-injection, but recovered completely within 1 month with the administration of valproate, lorazepam, olanzapine, and psychotherapy; Another patient experienced femur fracture during a physiotherapy session at 18 months post-injection.

Other AEs were classified as non-serious and only one was possibly related to the study. This was a patient (1/15) who experienced a first-ever partial motor seizure at month 6 (expected, possibly related to MS). As per the possible complications related to the long-term immunosuppressive treatment, one patient (1/15) developed a tremor during treatment with Tacrolimus which disappeared with dose-adjustment, one patient (1/15) developed respiratory failure, and infections were detected in three patients (3/15) (1 upper respiratory tract, 2 urinary tract). All the other reported AEs were related to non-study concomitant therapy or other medical conditions not related to the experimental protocol. The immunosuppressive treatment was successfully completed by patients. Tacrolimus was well tolerated by all patients except for the one already mentioned. In all patients, Tacrolimus blood levels were within the therapeutic target range (below 20 ng/mL).

As per the secondary objectives of the study, no changes were measured in EDSS and MSFC for the whole length of the study (**Table 4**).

**Table 4.**
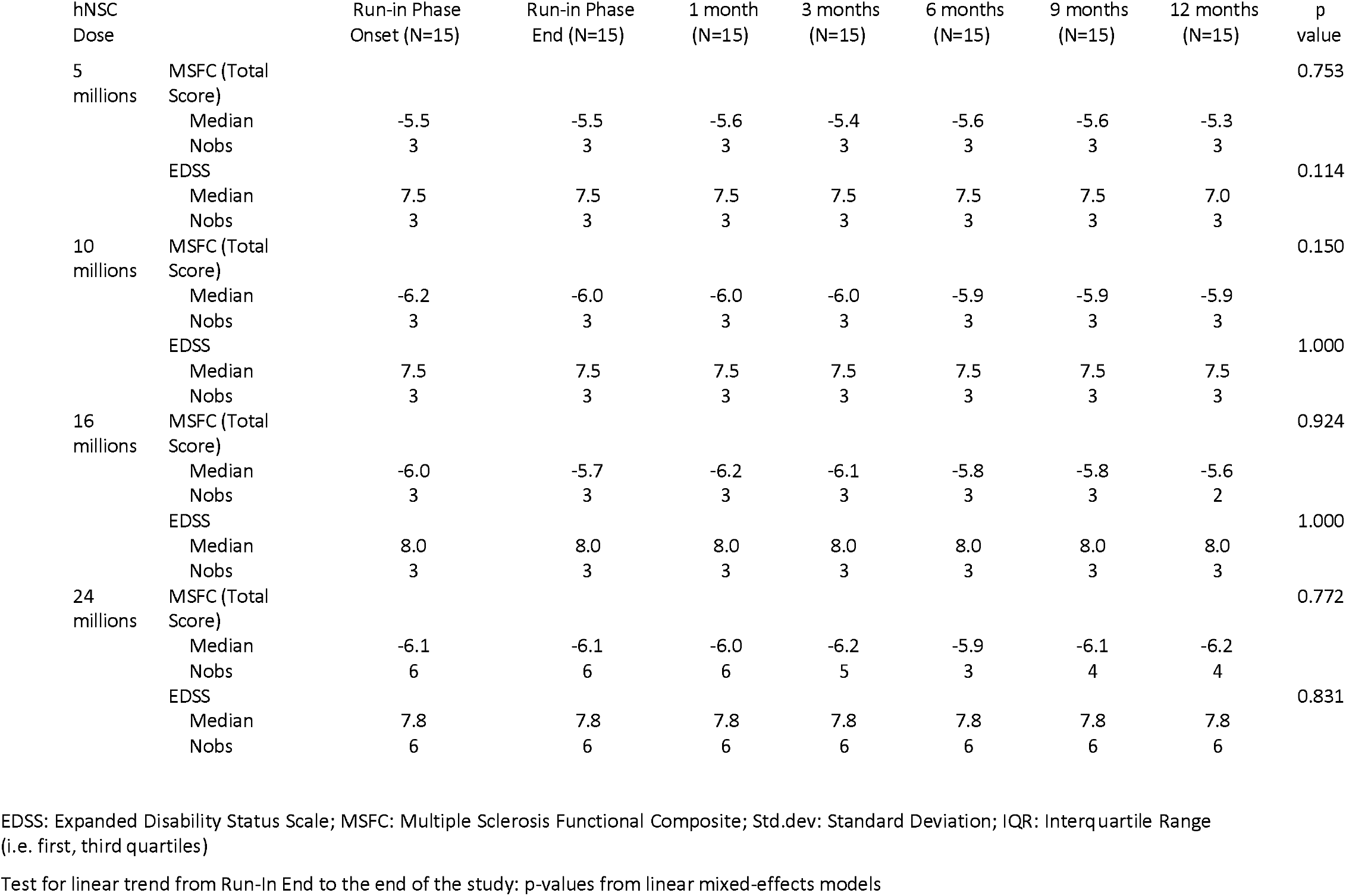
EDSS and MSFC evaluation

The median EDSS score did not change from the end of the run-in phase to the end of the study. Two patients (2/15) had a change in the functional systems score of >1 point, both in the pyramidal area: one decreased from 4.5 to 3.0 and another one increased from 0.0 to 2.0. The MSFC scores also did not significantly change from the end of the run-in phase to the end of the study.

None of the patients reported symptoms indicative of clinically active disease and cognitive functions, as measured by Rao’s BRB, did not show significant changes during the study period for any test. Neurophysiological parameters were monitored with EPs. Linear model analyses on ranks did not show any variation trends throughout the study for any visual, somatosensory, and motor EPs. We did not observe any changes on the OCT, except for one patient that showed an increase in retinal nerve fibre layer’s (RNFL) thickness in both eyes at month 6, which was interpreted as an artefact.

As part of the secondary objectives, of the 40 scheduled MRIs for the 15 patients, 25 were completed according to the study protocol and 15 were not performed (11 due to patient refusal, two due to patient inability, two due to the COVID-19 pandemic). One patient did not have a baseline MRI and post-Gadolinium images were not available for 8 MRIs due to exam interruption. Brain changes on MRI were classified as type 1, 2, and 3 to facilitate comparison between time points.

Type 1 changes were likely related to the surgical procedure. They included a linear T2-hyperintensity in the parenchyma beneath the surgical right frontal cranial hole, passing through the right frontal lobe white matter to the homolateral ventricle and corpus callosum (**Figure 1**).

**Figure 1.**
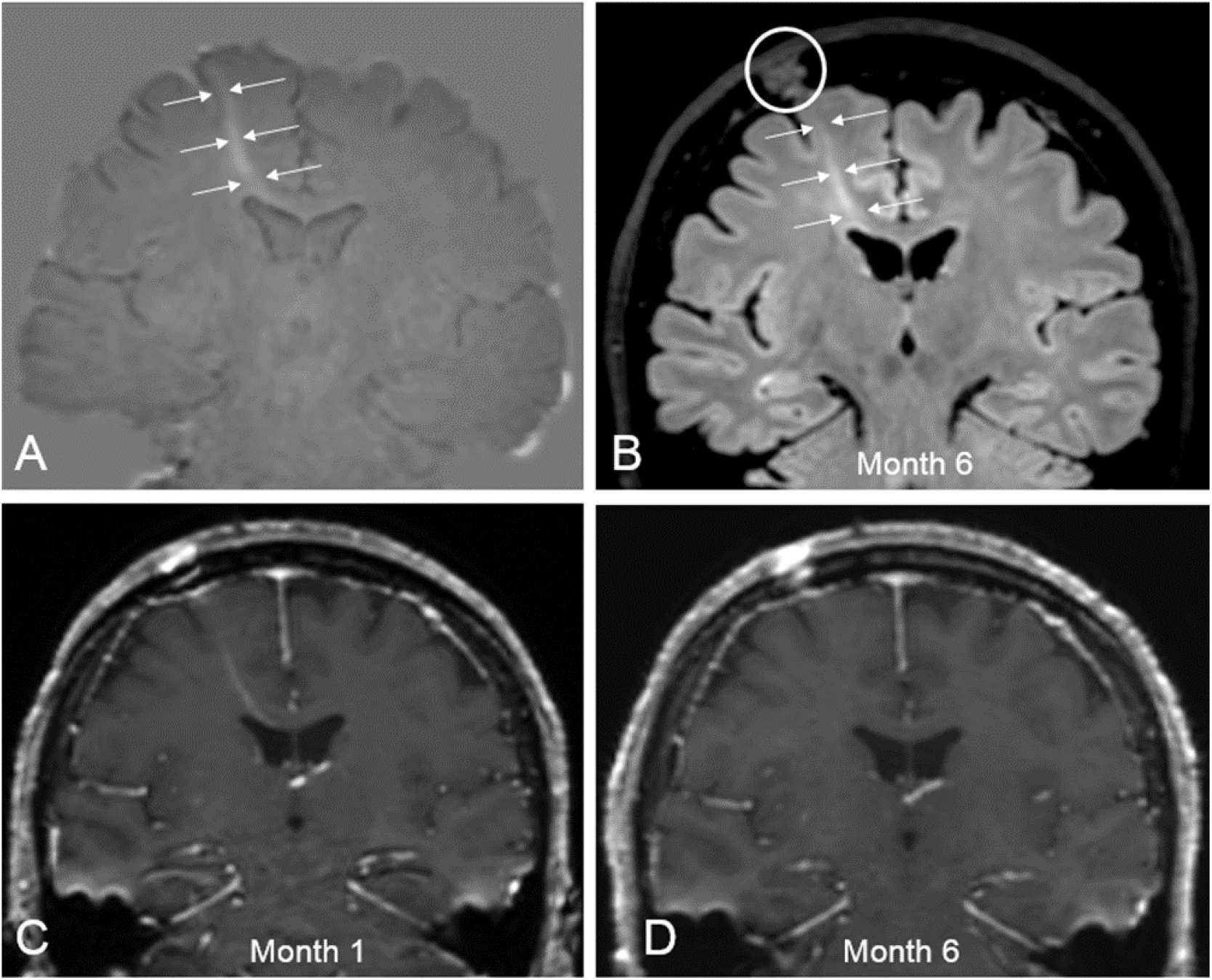
Example of type 1 change, interpreted as parenchymal gliotic modifications following ventricular cannulation. Coronal-oblique 3D-T2-FLAIR subtraction map in A (Month 12 minus Run-in End) detected a linearly-shaped positive signal change, passing through the right frontal lobe to the homolateral ventricle. This change corresponded to a subtle contrast-enhancing tract which occurred at Month 1 (B, post-Gadolinium T1-weighted image) at the level of the surgical cranial hole (circle in B). Month 6 (C) and Month 12 (D) T2-FLAIR images demonstrated the persisting chronic changes (arrows).

This was variable across all participants, ranging from a very subtle to a manifest signal change, and it was detectable in all patients on Month 1 images, together with a linear contrast enhancement on the corresponding post-Gadolinium T1-weighted images in most of cases.

Type 2 changes were MS-related and were used to compare the pre-transplantation with the post-transplantation period.

Type 3 changes were undetermined and detected in only one participant, where subtraction images highlighted a triangle-shaped, subtle T2-hyperintense signal change in the left frontal lobe white matter, without contrast enhancement or water diffusion restriction (**Figure 2**).

**Figure 2.**
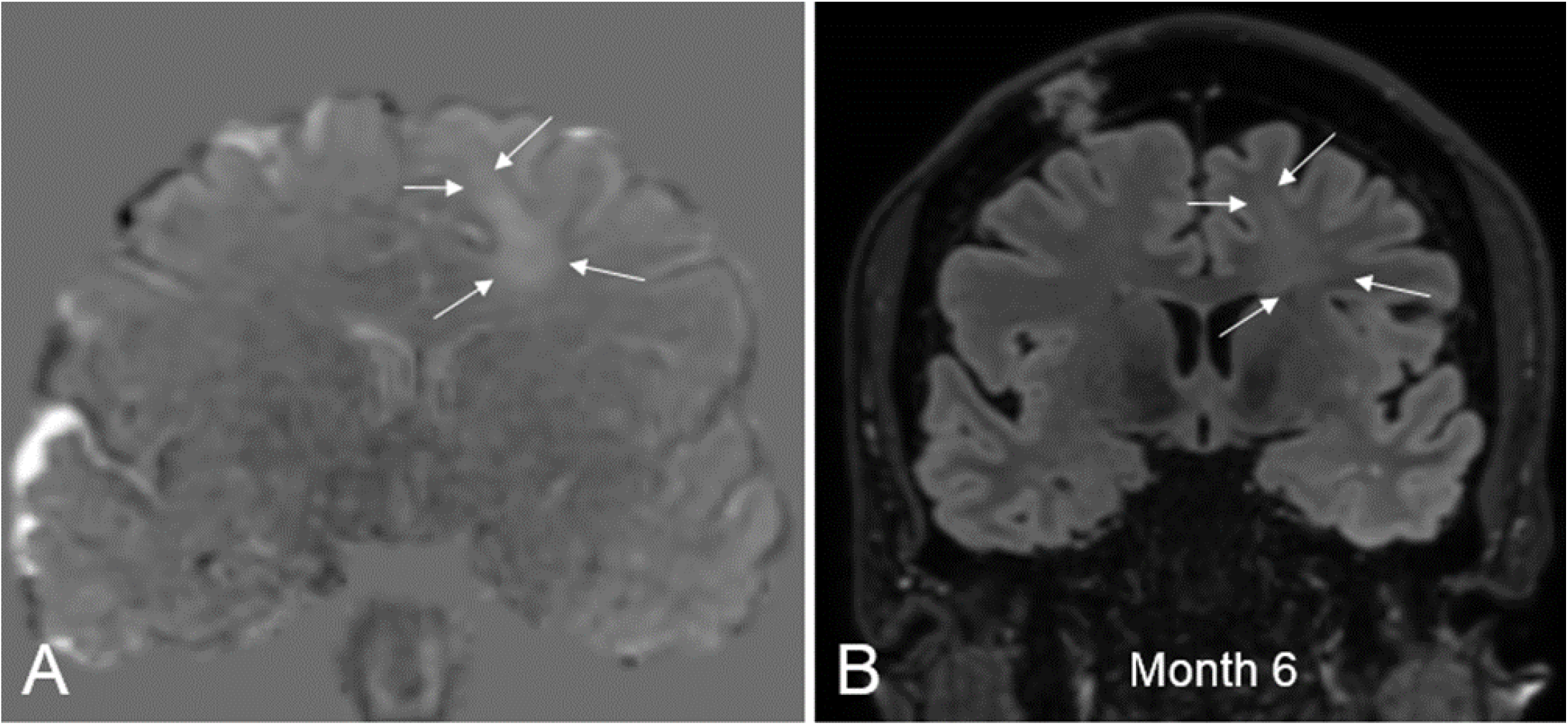
Type 3 changes. Coronal-oblique 3D-T2-FLAIR subtraction map (Month 6 minus Run-in-End) detects a triangle-shaped, positive signal change in the left frontal white matter (arrows). This corresponded to a subtle T2-FLAIR signal hyperintensity on the corresponding Month 6 source image (B, arrows). No contrast enhancement and/or water diffusivity restriction was noted (not shown).

This change remained stable at the subsequent follow-ups and was categorised as non-specific. This patient had additional MRI follow-up during which the lesion did not change.

Pre-transplantation (during the three-month run-in period), nine new-onset and two enlarging T2-visibile lesions were detected in 7/14 patients (50% of patients, with an average of 1.3 and 0.3 of new and enlarging T2 lesions per patient) (**Table 5**).

**Table 5.** Summary of pre- and post-transplantation new-onset and enlarging T2-visible lesions obtained by using subtraction imaging (see main text for details). N/A = data not available *When the Month12 time-point data was not available, the last previous available data was employed for subtraction

|  | **<u>Pre-Transplantation</u>** |  |  | **<u>Post-Transplantation</u>** |  |  |
| --- | --- | --- | --- | --- | --- | --- |
| **Subject #** | **Compared Time Points** | **Number of New (Enlarging) T2-visible lesions** | **Monthly Lesion Activity Rate** | **Compared Time Points\*** | **Number of New (Enlarging) T2-visible lesions** | **Monthly Lesion Activity Rate** |
| #1 | Run In End – Run In Onset | 0 | 0.00 | Month12 – Run In End | 8 | 0.67 |
| #2 |  | 0 | 0.00 | Month12 - Run In End | 1 | 0.08 |
| #3 |  | 0 | 0.00 | Month12 - Run In End | 0 | 0.00 |
| #4 |  | 2 | 0.67 | Month12 - Run In End | 6 | 0.50 |
| #5 |  | 1 | 0.33 | Month12 - Run In End | 0 | 0.00 |
| #6 |  | 0 | 0.00 | Month12 - Run In End | 1 | 0.08 |
| #7 |  | 0 | 0.00 | Month12 - Run In End | 0 | 0.00 |
| #8 |  | 0 | 0.00 | Month 6 - Run In End | 0 (1) | 0.17 |
| #9 |  | 0 (2) | 0.67 | Month12 - Run In End | 8 | 0.67 |
| #10 |  | 3 | 1 | Month12 - Run In End | 26 | 2.17 |
| #11 |  | 1 | 0.33 | Month12 - Run In End | 3 | 0.25 |
| #12 |  | 1 | 0.33 | Month 3 - Run In End | 0 | 0.00 |
| #13 |  | 0 | 0.00 | Month 9 - Run In End | 0 | 0.00 |
| #14 |  | N/A | N/A | Month12 - Run In End | 6 | 0.50 |
| #15 |  | 1 | 0.33 | Month12 - Run In End | 5 (2) | 0.58 |
| **Total** |  | **9 (2)** |  |  | **64 (3)** |  |

After transplantation (during the 12-month follow-up period), 64 new-onset and three enlarging T2-visible lesions were detected in 10 out of 15 patients (66% of patients, with an average of 6.4 and 0.3 of new and enlarging T2 lesions per patient). The annualised pre-transplantation rate of new or enlarging T2-visibile lesions was 3.30 (95% CI: 1·86–5·83), which was not statistically different (p-value= 0·26) to the post-transplantation rate (4·72, 95% CI: 2·32–9·61). When the 12-month data was not available, the last previously available data was used to compare).

Pre-transplantation, 10 lesions with contrast enhancement were detected in 6/14 patients (43%) (**Table 6**).

**Table 6.** Summary of pre- and post-transplantation lesions showing contrast enhancement on T1-weighted images, detected upon visual inspection. N/A = data not available.

| Subject # | Number of lesions with contrast enhancement (Time point) | Number of lesions with contrast enhancement (Time point) |
| --- | --- | --- |
| #1 | 0 | 1 (Month6) |
| #2 | 0 | 0 |
| #3 | 0 | 0 |
| #4 | 2 (Run in Onset) | 3 (Month3), 2 (Month6) |
| #5 | 1 (Run in End) | 0 |
| #6 | 0 | 0 |
| #7 | 0 | 0 |
| #8 | 0 | 1 (Month3) |
| #9 | 0 | 1 (Month6) |
| #10 | 1 (Run in Onset), 1 (Run in End) | 9 (Month3), 12 (Month 6), 4 (Month12) |
| #11 | 1 (Run in End) | 1 (Month9) |

|  |  |  |
| --- | --- | --- |
| **#12** | 1 (Run in Onset), 1 (Run in End) | 0 |
| **#13** | 0 | 0 |
| **#14** | N/A | 1 (Month12), 1(Month6) |
| **#15** | 1 (Run in Onset), 1 (Run in End) | 4 (Month3) |
| **Total** | **5 (Run in End), 5 (Run in Onset)** | **40** |

After transplantation, 40 gadolinium enhancing lesions were detected in multiple scans from 8 out of 15 patients. The annualised pre-transplantation rate of lesions with contrast enhancement was 2·86 (95% CI: 1·51–5·40) and it was not statistically different (p-value=0·98) to the post-transplantation values (2·83, 95% CI: 0·92–8·73).

Of note, 40% of the new-onset T2 lesions and the majority of gadolinium enhancing lesions happened in one single patient who had the highest disease activity in the run-in period. Additionally, one patient who showed one new T2 lesion and two lesions with contrast enhancement in the pre-transplantation period, had no new T2 or gadolinium enhancing lesions at follow-up.

Among the laboratory exams performed during the study, the only clinically significant variation was in the number of white blood cells that ranged from a mean of 7.4 x10^9/L at the end of the run-in period to a mean of 8.3 x10^9/L at the 12-month follow-up. The vital signs of the participants (blood pressure and temperature) were within the normal range and urine tests did not show clinically significant abnormalities. Biomarker analyses of NfL and CHI3L1/YKL-40 showed a statistically significant increase in the NfL concentration (pg/mL) at 1 month after intervention in both serum and CSF, followed by a gradual return to pre-intervention values (**Table 7**).

**Table 7.** Estimated marginal means, along with 95% confidence interval, from repeated measurements ANOVA models of serum neurofilament light concentrations (pg/mL) collected in NSC-SPMS patients at different groups and time points. P-values are referred to within-group and within-time comparisons and were derived from statistical contrasts defined into each model

| Biomarker | hNSC assigned dose groups | Run-In onset (RSTART) | Run-In end (REND) | 1 month (T1) | 3 months (T3) | 6 months (T6) | 9 months (T9) | 12 months (T12) | Within-group comparisons (p-values) |  |  |  |  |  |  |
| --- | --- | --- | --- | --- | --- | --- | --- | --- | --- | --- | --- | --- | --- | --- | --- |
|  |  |  |  |  |  |  |  |  | RSTART vs. REND | T1 vs. REND | T3 vs. REND | T6 vs. REND | T9 vs. REND | T12 vs. REND | p-value for trend |
| NFL CRMs | 5 millions | 13.84<br>(7.85-24.39) | 11.33<br>(6.43-19.97) | 31.61<br>(17.94-55.70) | 29.87<br>(16.95-52.63) | 20.62<br>(11.70-36.33) | 12.93<br>(7.34-22.78) | 13.21<br>(7.50-23.28) | 0.552 | 0.003 | 0.005 | 0.079 | 0.695 | 0.648 | 0.664 |
|  | 10 millions | 14.59<br>(8.28-25.71) | 16.91<br>(9.60-29.80) | 45.64<br>(25.90-80.43) | 21.78<br>(12.36-38.38) | 18.21<br>(10.33-32.08) | 17.17<br>(9.74-30.26) | 19.68<br>(11.17-34.68) | 0.660 | 0.004 | 0.452 | 0.826 | 0.964 | 0.652 | 0.825 |
|  | 16 millions | 11.80<br>(6.70-20.79) | 12.33<br>(7.00-21.73) | 98.03<br>(50.35-190.86) | 27.94<br>(15.85-49.23) | 23.07<br>(13.09-40.65) | 14.33<br>(8.13-25.25) | 14.84<br>(7.62-28.90) | 0.896 | <0.001 | 0.018 | 0.066 | 0.654 | 0.626 | 0.491 |
|  | 24 millions | 18.70<br>(12.53-27.91) | 14.90<br>(9.98-22.24) | 41.41<br>(27.74-61.81) | 26.05<br>(16.91-40.12) | 19.49<br>(11.41-33.29) | 16.18<br>(8.51-30.79) | 14.32<br>(8.91-23.00) | 0.340 | <0.001 | 0.030 | 0.370 | 0.814 | 0.882 | 0.540 |
|  | Overall | 14.53<br>(11.15-18.94) | 13.70<br>(10.51-17.86) | 49.19<br>(37.21-65.03) | 26.23<br>(20.06-34.29) | 20.27<br>(15.33-26.80) | 15.06<br>(11.23-20.20) | 15.33<br>(11.52-20.41) | 0.708 | <0.001 | <0.001 | 0.019 | 0.576 | 0.499 | 0.078 |
|  | Within-time comparisons (p-values) | 0.594 | 0.743 | 0.090 | 0.881 | 0.948 | 0.905 | 0.778 |  |  |  |  |  |  |  |
| GFAP | 5 millions | 77.40<br>(45.16-132.65) | 64.20<br>(37.46-110.02) | 79.41<br>(46.33-136.08) | 96.66<br>(56.40-165.65) | 83.13<br>(48.51-142.47) | 71.45<br>(41.69-122.45) | 81.26<br>(47.41-139.26) | 0.259 | 0.200 | 0.016 | 0.121 | 0.516 | 0.156 | 0.846 |
|  | 10 millions | 169.04<br>(98.64-289.70) | 144.04<br>(84.05-246.85) | 184.13<br>(107.44-315.55) | 144.11<br>(84.09-246.97) | 173.45<br>(101.21-297.26) | 191.89<br>(111.97-328.85) | 187.90<br>(109.64-322.01) | 0.333 | 0.140 | 0.998 | 0.262 | 0.086 | 0.111 | 0.469 |
|  | 16 millions | 148.40<br>(86.59-254.32) | 179.56<br>(104.77-307.73) | 222.80<br>(126.50-392.41) | 204.55<br>(119.35-350.55) | 176.34<br>(102.89-302.20) | 197.01<br>(114.96-337.64) | 180.73<br>(102.61-318.32) | 0.250 | 0.252 | 0.430 | 0.912 | 0.574 | 0.972 | 0.957 |
|  | 24 millions | 92.62<br>(63.28-135.57) | 97.47<br>(66.60-142.67) | 110.93<br>(75.79-162.36) | 124.85<br>(84.58-184.30) | 117.19<br>(77.00-178.37) | 110.82<br>(70.20-174.92) | 112.42<br>(75.25-167.96) | 0.661 | 0.269 | 0.049 | 0.215 | 0.461 | 0.287 | 0.301 |
|  | Overall | 115.80<br>(90.01-148.99) | 112.79<br>(87.67-145.11) | 137.87<br>(106.77-178.04) | 137.34<br>(106.66-176.83) | 131.39<br>(101.73-169.68) | 131.53<br>(101.45-170.54) | 132.72<br>(102.57-171.71) | 0.732 | 0.015 | 0.014 | 0.061 | 0.070 | 0.051 | 0.056 |
|  | Within-time comparisons (p-values) | 0.155 | 0.077 | 0.063 | 0.303 | 0.200 | 0.051 | 0.131 |  |  |  |  |  |  |  |

## DISCUSSION

The ICV injection of hNSCs was safe and did not lead to clinically relevant AEs post-intervention. The only SAEs that occurred were not related to hNSCs or the intervention, and no withdrawals or deaths were registered. Importantly, none of the patients has shown either a clinical relapse or a progression of their condition during the study period. The follow-up disability evaluation scales and cognitive testing documented clinical stability of all the participants. The neurophysiologic parameters and OCT also showed no significant variations. The MRI disease activity, assessed by annualised rate of new or enlarging T2-visible lesions and lesions with contrast enhancement, was not affected by the treatment. Finally, despite an initial fluctuation of the levels of NfL and CHI3L1/YKL-40, these markers later returned to their baseline values.

Currently, there is a lack of treatment options for SPMS. Although another study using a similar NSC preparation and method of administration is underway in another Italian centre (NCT03269071), to our knowledge this manuscript represents the first report of the use of hNSCs in SPMS patients. In addition, this study utilises a cell line that has been already shown safe in a prior clinical trial on ALS (Mazzini et al., 2015, 2019).

This study has limitations inherent to its early phase, non-randomised design, and small sample. Also, the fact that some MRIs could not be performed could have weakened the imaging results. Nevertheless, this research provides novel information on the safety and potential effectiveness of this treatment modality for SPMS. It also describes a system by which, owed to the peculiar expansion technique adopted for cGMP expansion (vescovi et al, 1997), the very same hNSCs from a single donor used here, can be used in a broad number of future clinical trials, thereby obviating to the current outstanding issues of inter-trial variability of cell drug products.

It is worth noting that, unlike other clinical trials, the cells used in this study are hNSCs that grow as stable, reproducible unmodified cell lines (**Table 8**)

**Table 8:** Comparison between primary culture (2014) and final product release test (2018 – 2020). Data for final product are expressed as mean value of the release tests conducted on all the batches used for the clinical trial. See Profico et al 2022 for complete quality control strategy. IP: intermediate product, FP: final product, CE: Clonal Efficiency; N: Neurons; O: Oligodendrocytes; A: Astrocytes; GC: Growth Curve, expressed as the slope of the growth curve..

|  | IP | FP |
| --- | --- | --- |
| CE | 1,70% | 1,45% |
| N | 27,90% | 31,50% |
| A | 36,10% | 50,50% |
| O | 17,70% | 18,70% |
| GC | 0,076 | 0,057 |

This is crucial in view of the emerging concept that one key issue in cell therapy is the inter-trial standardization of the cell drug, which may be homogeneous within the same clinical protocol but does vary significantly between different trials, due to the scarcity of the donor cells. The approach used here provides the significant advantage that the very same cGMP hNSCs used here - with all the patients receiving the same cell drug - will also be available for a number of future trials both on SPMS and other disorders. To the best of our knowledge, this is the first report of such an approach being possible.

In conclusion, NSC transplantation via ICV injection appears to be a safe procedure with neither major nor short-term deleterious effects. The study participants experienced a substantial clinical stability during 12 months of follow-up. The considerable absence of risks for the patients indicates a short-term neutral balance between benefits and risks and a therapeutic possibility on the horizon for SPMS patients. Further studies are needed to confirm and extend the findings herein and evaluate the actual therapeutic potential of advanced cell therapeutics for a condition where the lack of effective disease modifying therapies is a major unmet clinical need.

## Data Availability

All data produced in the present study are available upon reasonable request to the authors

## AUTHOR CONTRIBUTIONS

All the authors reviewed and approved the final manuscript

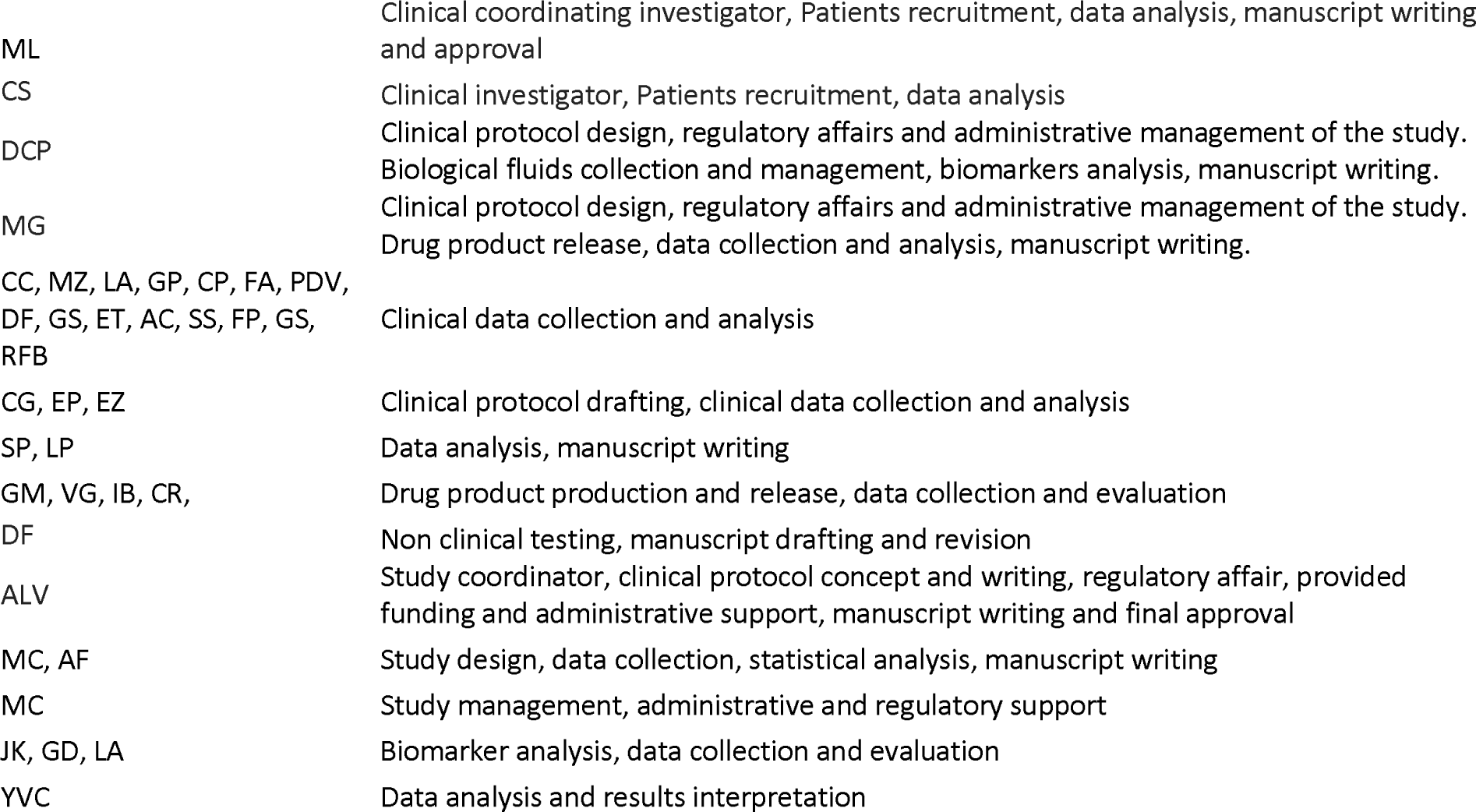

## DATA SHARING STATEMENT

All relevant data are included in this manuscript. The data that support the findings of this study are available from the corresponding authors upon reasonable request.

## ACKNOWLEDGEMENTS

Writing assistance in the preparation of this article was provided by Maria Carolina Rojido (Medical Writing Consultant). Proofreading and submission assistance was provided by Laura C Collada Ali (Medical Writing Consultant).

